# Return of Results in Psychiatric Genetics Research: Perceptions of Best Practices in a Global Sample of Psychiatric Genetics Researchers

**DOI:** 10.1101/2020.06.08.20125716

**Authors:** Gabriel Lázaro-Muñoz, Laura Torgerson, Stacey Pereira

## Abstract

Many research sponsors and genetic researchers agree that some medically relevant genetic findings should be offered to participants. The scarcity of research specific to returning genetic results related to psychiatric disorders hinders the ability to develop ethically-justified and empirically-informed guidelines for responsible return of results for these conditions. We surveyed 407 psychiatric genetics researchers from 39 countries to examine their perceptions of challenges to returning individual results and views about best practices for the process of offering and returning results. Most researchers believed that disclosure of results should be delayed if a patient-participant is experiencing significant psychiatric symptoms. Respondents felt that there is little research on the impact of returning results to participants with psychiatric disorders and agreed that return of psychiatric genetics results to patient-participants may lead to discrimination by insurance companies or other third parties. Almost half of researchers believed results should be returned through a participant’s treating psychiatrist, but many felt that clinicians lack knowledge about how to manage genetic research results. Most researchers thought results should be disclosed by genetic counselors or medical geneticists and in person, however, almost half also supported disclosure via telemedicine. This is the first global survey to examine the perspectives of researchers with experience working with this patient population and with these conditions. Their perspectives can help inform the development of much-needed guidelines to promote responsible return of results related to psychiatric conditions to patients with psychiatric disorders.

## INTRODUCTION

Guidelines for responsible return of individual genetic research results should be context dependent^1,2,3,4,5^ to ensure that return maximizes net benefit and is responsive to the needs and characteristics of the participant populations. Given recent advances in identifying genomic correlates of polygenic conditions, such as psychiatric disorders, and the expanded use of more comprehensive single-nucleotide polymorphism (SNP) arrays and genome and exome sequencing, psychiatric genetics researchers are increasingly managing questions about whether and how to return individual results to participants.^6^ There is an emerging consensus among genetics researchers, including psychiatric geneticists,^7,8,9,10^ and research sponsors that some medically relevant genetic findings should be offered to participants. In addition, most research participants expect that researchers will return medically relevant information to them.^11,12^

Given the objective and methodological approaches (e.g., genome-wide association studies (GWAS) comparing cases and controls) in psychiatric genetics research, a substantial portion of the participant population will have a diagnosis of at least one psychiatric disorder. An important consideration in this research and in the development of guidelines for returning individual research results is that, depending on the psychiatric disorder examined, patient-participants in these studies are more likely than control-participants or participants in many other genomics research fields to have cognitive impairments or pathological emotional processing and responses. This may increase the likelihood of participants misunderstanding the implications of results or having a negative emotional response. There is, however, a dearth of research about the impact of returning results to individuals at risk for psychiatric disorders or who have a psychiatric diagnosis.^13^ One of the few relevant studies found that individuals with depressive symptoms who were told that they were at an increased genetic risk for depression were more likely to believe they were currently experiencing major depression or would experience it in the future (“prognostic pessimism”) compared to people with depressive symptoms told they were not at an increased genetic risk for depression.^14^

More research has been done on the psychological and psychosocial impacts of disclosing genetic information to “healthy” individuals (participants from the general population) or individuals with other health conditions, such as cancer or heart disease. Most of these studies have measured anxiety, depression, or other symptoms of psychological distress in response to receiving predictive genetic information and have typically not found evidence of significant or sustained negative psychological effects.^15,16,17^ Other studies, however, show that many individuals with known disease risk, such as Huntington’s disease or some types of cancers, often decline or delay testing, and some data suggest that those less willing to get tested are those more likely to believe they will feel psychological distress if they learn they are at risk.^15,18,19^ Thus, those who may be more prone to psychological distress in response to receiving genetic results may be less likely to enroll in studies that have assessed those harms. Additionally, there has been debate about whether quantitative measure of psychological distress, on which many of these studies have relied for drawing their conclusions, are the best way to assess the emotional impact of this information.^20^ Further, several studies have reported other negative impacts of receiving genetic information. For example, in one study, participants from the general population who were told they were at an increased genetic risk for developing depression using a sham genetic test reported more depression symptoms over the previous two weeks than those told they did not have such genetic predisposition.^21,22^ In another study, healthy older adults who knew that they were at increased genetic risk for Alzheimer’s disease (i.e., carriers of the *APOE4* allele) judged their memory as worse on subjective memory scales and performed worse on an objective memory test than individuals who also carry the *APOE4* allele, but were unaware.^23^

Empirically-informed and ethically-justified guidelines for returning genetic research results to patient-participants in psychiatric genetics research are critically needed because this is a quickly expanding field of genetics and return of results is a growing practice in this area (GLM, unpublished data).^7,8^ Furthermore, though there is a lack of data on how patients with psychiatric disorders actually respond to genetic research results, the symptoms that characterize many psychiatric disorders suggest these patients are more likely to experience psychosocial harms in response to receiving results compared to control or “healthy” participants. Additionally, some research has found transient psychosocial impacts of returning genetic findings even in individuals with no psychiatric disorders.^24^ To develop these guidelines, the psychiatric genetics researchers’ perspectives about how to return results to these patients are essential; these researchers have both the relevant knowledge of what types of results may be discovered, as well as experience with this population. Thus, we conducted a survey examining these and related issues in a global sample of psychiatric genetics researchers.

## MATERIALS AND METHODS

### Participant Sampling

Members of the International Society of Psychiatric Genetics (ISPG), the largest international society of psychiatric geneticists, and attendees of ISPG’s 2019 World Congress of Psychiatric Genetics (WCPG) were invited via email or in person to participate in a web-based survey between July 2019 and December 2019. The Institutional Review Board (IRB) at Baylor College of Medicine approved the study. For those invited via email, reminders were sent up to three times. To increase our response rate, participants were offered a chance to win one of six $200 gift cards.

### Survey Measures

The survey was developed based on the extant literature and the results of a previous study in which we (GLM, SP) interviewed 39 psychiatric genetics researchers from 17 countries about their perspectives toward returning genetic research results to individual participants.^7,8^ Except for preferred professional and preferred modality for results disclosure, all data were collected with five-point Likert items with response options from “strongly disagree” to “strongly agree” with a neutral midpoint (“neither agree nor disagree”). Preferred professional and modality for returning results were queried by asking respondents to choose their preference from a list of seven professionals and modalities. Survey instructions stated that questions were about adult case-participants as opposed to adult control-participants, unless otherwise specified, and that adult case-participants would be referred to as patient-participants.

A social scientist (SP) conducted two cognitive interviews^25^ with psychiatric genetics researchers to assess question relevance, readability, face validity, comprehension, and survey length, which led to minor changes. The survey was then tested by 10 colleagues who are not in the psychiatric genetics field and piloted with five psychiatric genetics researchers. No changes were necessary based on the pilot. The survey was distributed via Qualtrics and took approximately 15-20 minutes to complete.

### Data Analysis

We report response frequencies for each item. Likert item data are reported as agree, disagree, and neither agree nor disagree by combining the two responses on each end (e.g., combining strongly agree and agree). For preferred professional and modality for disclosure of results, we report the percentage of respondents that selected each option. Differences in sample sizes reflect missing responses.

## RESULTS

### Participant Characteristics

We invited 2,024 psychiatric genetics researchers to participate in the survey; 490 individuals opened the link. Nine people indicated they did not want to participate. Of the 481 people who agreed to participate, 74 did not provide answers to any questions, leaving 407 respondents (85%) for analysis. Our final response rate was 20.1% of those invited. Participant demographics are reported in Table 1. We received responses from researchers from 39 different countries. Approximately half (54%) of researchers were female, 28% held MDs, and 58% held a PhD without an MD degree. Sixty-six percent reported they were responsible for “overall study design” and 81% were involved in analysis of genomic samples/data. The majority (86%) reported they used array-based testing (e.g., SNP arrays) in their research and many were also using genome (48%) and exome (38%) sequencing and single-gene testing (32%). Respondents’ roles, type of genetic testing used, disorders examined, and patient populations are shown in Table 2.

**Table 1.** Psychiatric Genetics Researchers’ Demographics.

|  | **% (n)** |
| --- | --- |
| **Total** | 100% (407) |
| **Gender (n=350)** |  |
| Female | 54% (189) |
| Male | 43% (150) |
| I prefer not to say | 3% (11) |
| **Country (n=334)<sup>1</sup>** |  |
| United States | 42% (139) |
| United Kingdom | 10% (32) |
| Canada | 9% (30) |
| Germany | 4% (14) |
| Brazil | 4% (13) |
| Norway | 4% (13) |
| Australia | 3% (11) |
| Sweden | 3% (11) |
| Other European Countries | 15% (51) |
| Asian Countries | 5% (18) |
| Other Countries in the Americas | 4% (12) |
| African Countries | 2% (7) |
| Other Oceania Countries | .3% (1) |
| **Academic degree (n=351)<sup>1</sup>** |  |
| Ph.D. only | 56% (202) |
| M.D. only | 15% (51) |
| M.D. and Ph.D. | 13% (46) |
| M.S., Genetic Counseling only | 3% (10) |
| Other | 12% (42) |
| **Years in psychiatric genetics research (n=343)** |  |
| 0-4 years | 25% (85) |
| 5-9 years | 33% (114) |
| 10-19 years | 26% (88) |
| 20-29 years | 11% (37) |
| 30+ years | 5% (19) |

<sup>1</sup>Respondents could select all responses that applied.

**Table 2.** Psychiatric Genetics Researchers’ Roles, Testing, and Populations.

|  | **% (n)** |
| --- | --- |
| **Role (n=407)<sup>1</sup>** |  |
| Analysis of genomic samples/data | 82% (332) |
| Overall study design | 66% (270) |
| Collection of clinical data and biospecimens | 38% (154) |
| Generating genomic data | 34% (138) |
| Obtaining informed consent | 26% (104) |
| Providing clinical care | 18% (74) |
| Other | 4% (18) |
| **Genetic Test Used in Research (n=405)<sup>1</sup>** |  |
| Array-based testing (e.g., SNP arrays) | 86% (348) |
| Genome sequencing | 48% (195) |
| Exome sequencing | 37% (152) |
| Single-gene testing | 32% (131) |
| Panel-based testing | 14% (55) |
| Karyotyping | 6% (26) |
| Other | 6% (24) |
| **Psychiatric Disorder Studied (n=352)<sup>1</sup>** |  |
| Schizophrenia and Related Disorders | 55% (193) |
| Depressive Disorders | 41% (146) |
| Bipolar Disorder | 38% (135) |
| Autism Spectrum Disorder | 25% (89) |
| Attention-Deficit Hyperactivity Disorder | 19% (67) |
| Anxiety Disorders | 16% (57) |
| Obsessive-Compulsive Disorder and Related | 12% (42) |
| Alzheimer's Disease | 11% (38) |
| Eating Disorders | 10% (36) |
| Substance Abuse / Addiction | 9% (31) |
| Tourette's Syndrome | 6% (20) |
| Suicide | 2% (7) |
| Huntington's Disease | 2% (6) |
| Other | 9% (32) |
| **Patient Population (n=405)<sup>1</sup>** |  |
| Adults | 94% (382) |
| Children | 44% (179) |
| Adults lacking decision-making capacity | 14% (58) |
| Children not expected to have decision-making capacity as adults | 12% (49) |

<sup>1</sup>Respondents could select all responses that applied.

### Challenges to Responsible Return of Results in Psychiatric Genetics Research

Respondents’ perspectives toward challenges to offering to return individual genetic research results related to psychiatric disorders to patient-participants are reported in Figure 1. Most researchers (77%) felt that a significant challenge to offering return of results is that patient-participants could have a negative emotional reaction in response to receiving results, and that little research exists about the impact of returning results to patient-participants (75%). Furthermore, nearly half (48%) of respondents agreed return of results should be delayed if a participant is experiencing significant psychiatric symptoms. The vast majority of researchers (89%) also agreed that patient-participants may misinterpret or misunderstand results. On the other hand, most researchers agreed that practices for returning medically relevant findings should be the same for patient-participants and controls (66%), and the same for results related to psychiatric disorders and non-psychiatric disorders (66%). Most agreed that other significant challenges to returning results are that clinicians lack knowledge and understanding about how to manage results (78%), results generally lack individual-level meaning (72%), and results often lack implications for treatment (83%).

**Figure 1.**
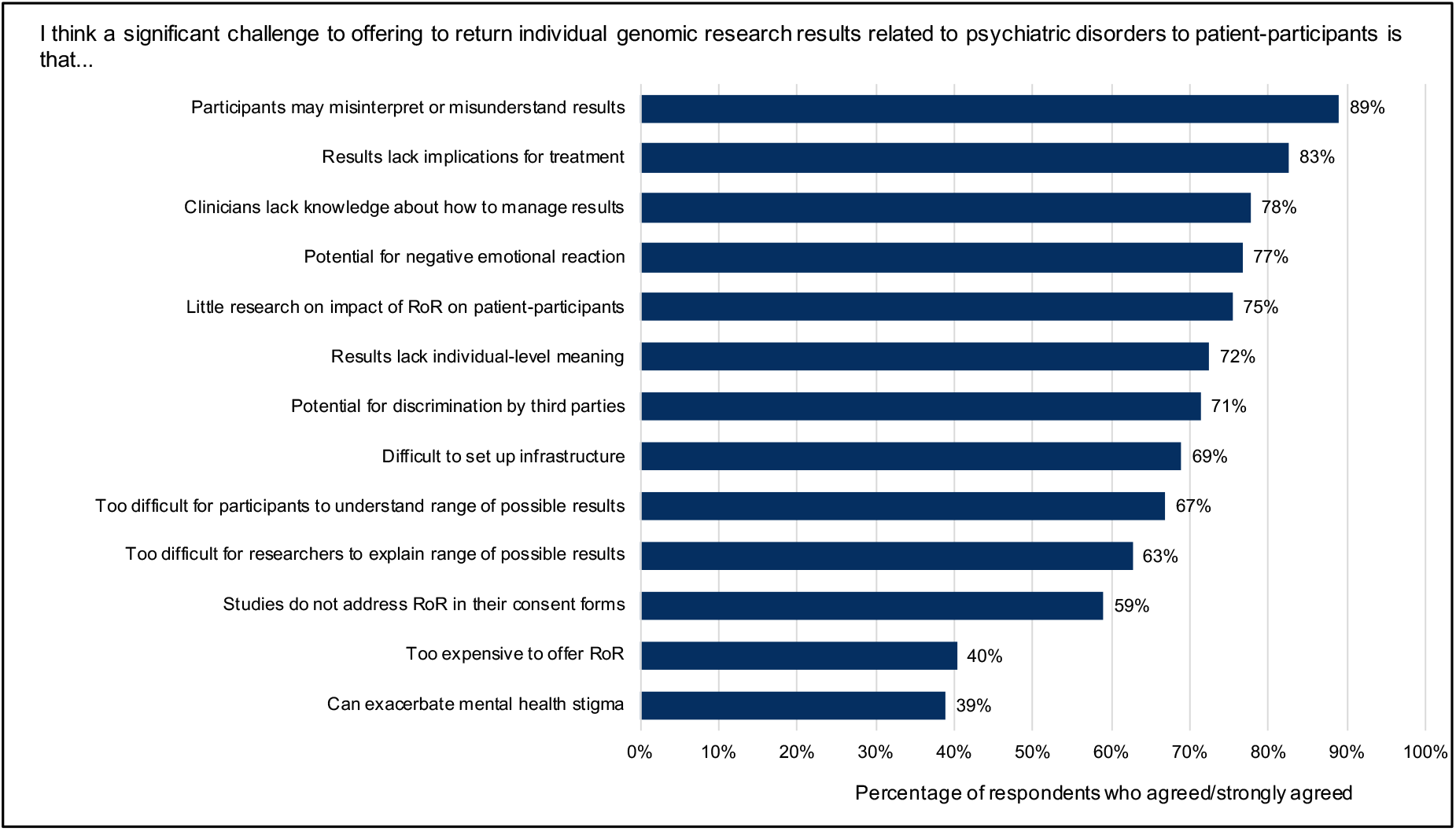
Psychiatric Genetic Researchers’ Perspectives on Challenges to Offering to Return Results Related to Psychiatric Disorders to Patient-Participants. Response options ranged from *strongly disagree* to *strongly agree*, with a neutral midpoint. Percentage of respondents choosing *agree* or *strongly agree* is shown for each challenge.

Researchers also noted practical and societal challenges. Most (59%) agreed that a challenge to returning results is that many studies do not address return of results in their consent forms. In fact, only 40% of respondents indicated that their own studies’ consent forms addressed the issue of whether results would be returned to participants, with an additional 15% addressing it only in some of their studies’ consent forms. Most agreed that when obtaining informed consent, it is too difficult for researchers to explain to participants the range of results that could be generated in the study (63%), as well as too difficult for participants to understand that range (67%). A substantial number agreed that it is difficult for researchers to set up the infrastructure necessary to return results (69%), and that it is too expensive to offer return of results (40%). Finally, 39% of researchers agreed that a significant challenge to returning results related to psychiatric disorders is that it could exacerbate mental health stigma, and that it could lead to discrimination by insurance companies and/or other third parties, such as schools and banks (71%).

### Perceptions of Best Practices When Offering Return of Results to Patient-Participants

Researchers were asked about the process of offering individual genetic research results related to psychiatric disorders to patient-participants. Most (84%) agreed that patient-participants should be able to opt out of receiving all results related to psychiatric disorders, and 87% agreed that participants should be able to opt out of receiving specific types of results related to psychiatric disorders (e.g., medically actionable vs. non-medically actionable). Respondents (71%) also felt that participants should be able to choose whether research results related to psychiatric disorders are included in their medical records. Finally, a third of researchers (34%) felt that when using genome/exome sequencing, psychiatric genetics researchers have a responsibility to look for medically actionable information (e.g., ACMG-59) even when it is not the focus of the study, and 32% agreed that psychiatric genetics researchers have a responsibility to reanalyze genomic data over time and recontact participants if medically relevant findings are discovered. Forty percent of respondents agreed that researchers have a responsibility to offer results related to psychiatric disorders discovered incidentally, but many were ambivalent about this, with 33% selecting neither agree nor disagree.

### Perceptions of Best Practices about the Process of Returning Results

Most researchers (71%) agreed that results related to psychiatric disorders should be confirmed by a clinically certified laboratory before being returned to participants. Many respondents were unsure or ambivalent about to whom the results should be disclosed. When asked whether results related to psychiatric disorders should be returned *directly* to participants (or their legal guardian, if applicable), a third of respondents agreed, a third disagreed, and a third selected “neither agree nor disagree.” On the other hand, 41% agreed that results should be returned *indirectly* through a participant’s treating psychiatrist, and a third of respondents selected “neither agree nor disagree.”

Participants were also asked by whom and via what modality they thought medically relevant genetic research results related to psychiatric disorders should be disclosed to participants. Respondents were most supportive of results being returned by those with clinical genetics expertise, including a genetic counselor (89% agreed) and a medical geneticist (77%). They were less supportive of results being returned by the patient’s treating psychiatrist (57%), a physician researcher (36%), the patient’s primary physician (25%), or a non-clinician researcher (11%). When asked which type of professional would be their *preferred* person to return findings, the majority (53%) selected genetic counselor and 20% selected medical geneticist (Figure 2).

**Figure 2.**
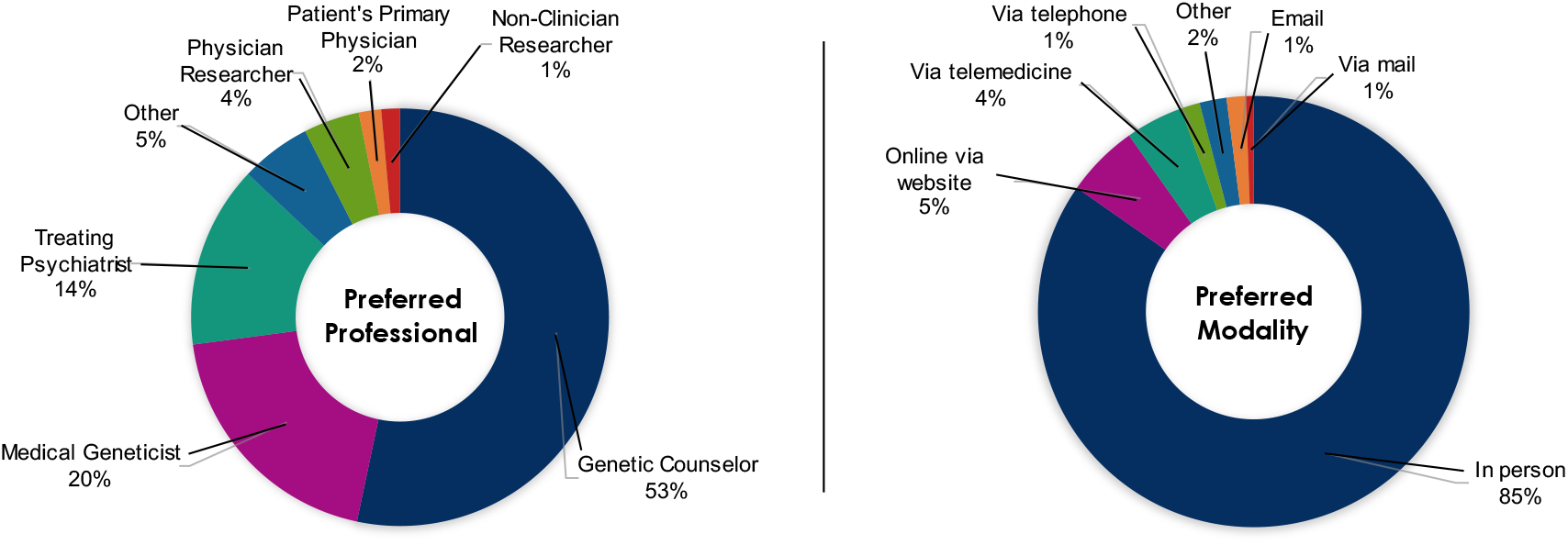
Preferred Professional and Modality to Return Medically Relevant Genomic Research Results Related to Psychiatric Disorders to Patient-Participants. Researchers were asked to choose which professional (left) and which modality (right) was their *preferred* for returning medically relevant genomic research results related to psychiatric disorders from a list of 7 options for each. Percentage of respondents who chose each option are shown.

When asked via which modality they thought medically relevant genetic research results related to psychiatric disorders should be returned, the vast majority of respondents (92%) agreed that such results should be returned in person, while 45% agreed they should be returned via telemedicine (secure video call). Researchers were less supportive of other options: 21% agreed that results should be returned online via a secure website, 17% via telephone, 9% via mail, and 7% via email. When asked to choose which modality would be their *preferred* method for returning results, 85% selected in person and far fewer respondents selected each of the remaining options (Figure 2).

## DISCUSSION

### Concerns about Psychosocial Impacts

Respondents agreed throughout the survey that there is potential for patient-participants to experience negative psychosocial impacts in response to receiving genetic research results related to psychiatric disorders. Most researchers believed patient-participants could have negative emotional reactions to results, may misinterpret results, and that there is little research on the impact of returning findings to these participants. This is consistent with our previous findings in which psychiatric genetics researchers expressed concern about how patient-participants’ “cognitive function may not be as good as other people’s, so it [could] be very easy to misread the information […]” and how in this research “we have highly anxious people, we have depressive people. They consistently take information more negatively that they should.”^7^ In line with these findings, researchers believed that return of results should be delayed if a participant is experiencing significant psychiatric symptoms. When asked whether return of results practices should be different for patient-participants and control-participants, however, researchers disagreed. This may be explained by psychiatric genetics researchers’ concerns about the perpetuation of stigma and undue discrimination that patients with psychiatric disorders often experience.^**Error! Bookmark not defined**.,26^ Most researchers agreed that returning results could lead to discrimination by insurance providers, schools, banks, and other third parties. This could be a reason why most researchers also thought participants should be able to choose whether medically relevant findings should be included in their medical records. Once in the medical record, it becomes easier for insurance providers (e.g., life insurance providers) and other third parties to gain access to this information.^27,28,29^ Thus, there is tension between wanting to return results that are medically relevant, but also wanting to protect these patients from psychosocial harms and potential discrimination.

### Structural Challenges

#### Consent

Respondents agreed there are a number of practical and structural challenges to responsibly returning psychiatric genetic research results to patient-participants. First, most agreed that one such challenge is that many studies do not address return of results in their consent forms. Guidelines and best practice standards recommend that the issue of return of results be addressed in genetic research consent forms, and many recommend that research participants be given the option whether they want to receive individual research results at the time of enrollment before such results are discovered.^3,9,30,31,32,33^ In fact, some recommend that results *not* be returned if the participant has not previously consented to receiving this information. Much genetic research, including psychiatric genetic research, however, is conducted using biospecimens or data from biospecimens that were collected before the possibility of widespread return of individual research results was anticipated. As such, the consent forms with which these biospecimens and data were obtained may not have addressed the issue of return of results or may have explicitly stated that results would not be returned. Many of our respondents reported that their consent forms often did not address return of results, or they were unaware of whether the issue was addressed. This is line with other studies that have found that the majority of genetic research consent forms either stated explicitly that genetic results would <u>not</u> be returned or did not address the issue at all.^34,35^ Further, respondents agreed that it is difficult when obtaining consent for researchers to explain and for participants to understand the range of results that could be generated in these studies. This highlights the need for more research in this area about how potential individual genetic research results could be best communicated to participants.

#### Lack of Infrastructure

A second practical challenge to returning results that our respondents confirmed was the lack of infrastructure and resources necessary to do so responsibly. These challenges have been a common refrain among researchers and other stakeholders in genetics research alongside the growing consensus to return some medically relevant results to participants. Most laboratories may not have the resources or experience contacting participants to return results in a way that minimizes potential emotional harm and ensures participants understand the implications of the findings, a concern our respondents noted. In fact, most laboratories likely do not have clinicians qualified to disclose this information. Furthermore, previous research suggests that many psychiatric genetics researchers believe it is important to help ensure that patient-participants whose results reveal or confirm increased risk for a psychiatric disorder have access to care.^8^ The lack of clinicians that could communicate the implications of findings on many projects and concern about follow-up care may explain why an unexpectedly high number of respondents agreed that patient-participant results should be returned through the participant’s treating psychiatrist. Interestingly, even if the results were returned directly to the treating psychiatrists, most researchers believed that a significant challenge is that clinicians lack knowledge and understanding of how to manage results. Finally, most respondents agreed that research results should be confirmed by a clinically certified laboratory before returning them to participants, which represents additional logistical and financial burdens.

Establishing an infrastructure for returning results to participants that meets researchers’ ideal for how this should be done would require significant investments from research sponsors. Some have expressed concern that this could divert funds away from research.^36^ However, if research sponsors provide the resources necessary to develop this infrastructure, returning results would be a way to demonstrate reciprocity for patients’ participation by providing them with information they want and that could benefit their health. Furthermore, it could increase the societal benefit of investing in psychiatric genetics research and may incentivize participation as many researchers in this field believe (GLM, unpublished data)^7^ and research has shown.^37,38,39,40^ Thus, as psychiatric genetics knowledge grows, some patient-participants could benefit directly from these research efforts.

### Perceptions of Best Practices when Offering Results

The American College of Medical Genetics and Genomics (ACMG) recommendation that laboratories should analyze and report “incidental” or secondary findings when conducting genome or exome sequencing in clinical settings generated considerable debate about whether researchers should follow similar practices.^41,42,9^ An influential article by Jarvik and colleagues argued that researchers have a responsibility to offer to return medically actionable findings and may be ethically and scientifically justified in offering some non-medically actionable findings, but that participants should be able to opt out of receiving any findings.^9^ Psychiatric genetics researchers seem to support this notion. In previous publications, we have reported that the vast majority of psychiatric genetics researchers agree medically actionable findings should be offered to participants, and here we found that most researchers agree participants should have the opportunity to opt out of the return of results and even opt out of the return of specific categories of results (e.g., medically actionable vs non-medically actionable) (GLM, unpublished data).^7,8^

Jarvik and colleagues’ article, however, maintained that researchers do not have a duty to analyze and offer findings that are not within the scope of the research (“duty to hunt”).^5,9^ Interestingly, a third of researchers in our sample agreed that they do have a responsibility to look for medically actionable information (e.g., ACMG-59) even when it is not the focus of the study. A substantial number of researchers also agreed that they should offer to return results discovered incidentally. Furthermore, there has been significant debate about whether researchers and clinicians have a responsibility to reanalyze genomic data and recontact patients or participants if the interpretation of a genomic finding changes in a way that could have medical implications.^3,43,44^ The general consensus has been that researchers do not have a duty to reanalyze in part due to feasibility constraints once a study’s funding has ended;^3^ however, the American Society of Human Genetics and others have recently expressed support for a limited duty to recontact in the research context.^45^ About a third of researchers in our sample agreed that researchers should reanalyze genomic data over time and recontact participants if medically relevant findings are discovered.

### Perceptions of Best Practices about the Process of Returning Results

Respondents felt that if medically relevant genetic research results were to be returned to participants, they should be disclosed by a clinical genetics professional, with most supporting disclosure by genetic counselors. This is consistent with recommendations and research that urges disclosure of genetic research results by a professional who has expertise in both genetics and communication of such information.^30,46,47^ Though this may be the ideal, high costs and shortages of genetic counselors, particularly in some areas of the world, may make this a non-scalable solution for now. Further, our respondents were most supportive of returning results in person, which is often noted as ideal yet unrealistic due to issues of limited workforce, efficiency, and cost.^48^ This is also impracticable for those living outside urban settings, where most genetic services are offered,^49^ or in other areas of the world where genetics specialists are in short supply. While studies have found telephone delivery of genetic results to be a tenable alternative to in-person disclosure,^3,50^ our respondents were not very supportive of this mode of delivery. In order to meet demand as returning individual results to participants becomes increasingly common, however, other, more scalable options will be necessary. Our respondents were more supportive of the use of telemedicine over telephone for returning results, which may reduce burden on both the research and participant side, and therefore maximize the capacity of relevant genetic specialists to return results. It is also important to note that these data were collected before the COVID-19 pandemic, during which many integrated teleconferencing into medical and research practices. This may lead to more acceptance of telemedicine. Some challenges will remain, though, including issues around access, privacy, and providing services across jurisdictions.

### Limitations

We sampled a diverse group of psychiatric researchers across 39 countries, but results may not be representative of the larger population of psychiatric genetics researchers. Because respondents self-selected for participation, it is possible that those with stronger opinions or those who were more familiar with the issue of return of research results may have been more likely to respond. There is also potential for social desirability bias with some survey questions due to the aforementioned emerging consensus in the field that some medically relevant research results should be offered to participants. Notwithstanding, this is the first study to assess the perspectives of an international sample of psychiatric genetics researchers on the challenges of and ideal practices for returning results to their participant populations.

## CONCLUSION

Guidelines for safe and responsible return of genetic research results to participants should be context specific. Our findings indicate that many researchers feel that the potential for patient-participants in psychiatric genetics studies to have a negative emotional response or misunderstand results are significant challenges to returning results in this field and that return of results should be delayed if a participant is experiencing significant psychiatric symptoms. Respondents also agreed that there are a number of practical and societal challenges. Though respondents felt that genetic results should ideally be disclosed by a genetic counselor and in person, they were moderately supportive of other options that may be more scalable, such as telemedicine. Given recent advances in psychiatric genetics research alongside an emerging international consensus that some medically relevant genetic research findings should be offered to participants, guidance on how to responsibly return results to this population is critically needed. Future research should explore options for maximizing benefit and minimizing harms to psychiatric genetics patient-participants, while exploring scalable solutions for returning individual research results.

## Data Availability

The datasets generated during and/or analysed during the current study are available from the corresponding author on reasonable request.

## ACKNOWLEDGEMENTS

Research for this article was funded by the National Human Genome Research Institute (NHGRI) of the National Institutes of Health (NIH) Grant R00HG008689 (Lázaro-Muñoz, G). The views expressed are those of the authors alone, and do not necessarily reflect views of NIH.

